# Postpartum hemorrhage in a large prospective cohort study: is uterine atony really the main culprit?

**DOI:** 10.64898/2026.01.27.26344932

**Authors:** Wolfgang Korte, Torsten Hothorn, Justus Bürgi, Matthias Rösslein, Nicole Ochsenbein, Christian Haslinger

## Abstract

**Background:** Uterine atony (∼70%), lacerations (∼20%) and placenta-related problems (∼10%) are assumed main reasons for postpartum hemorrhage genesis. Coagulation components predictive for postpartum blood loss can be identified prepartum and before traditionally assumed main reasons are observed.

**Objectives:** To better understand postpartum hemorrhage genesis, we prospectively researched prepartum clinical information, presence of assumed main reasons and peripartum coagulation changes in parturient women.

**Study design:** In 676 women with vaginal deliveries, age, BMI, parity, gestation age, duration of second stage of labor and presence and type of assumed main reasons (uterine atony, lacerations and placenta-related problems) were recorded. Measured blood loss within 24h postpartum defined no, non-severe or severe PPH (<500ml, ≥500ml to <1000ml, ≥1000ml). Hemoglobin, platelet count, fibrinogen, factor II and factor XIII activity were measured at admission and 24-48h postpartum.

**Results:** Of 191 women developing postpartum hemorrhage, 53.9% did not show assumed main reasons (expected <5%, p<.001). Of 45 women with severe postpartum hemorrhage, 15.5% were without assumed main reasons (<5%, p<.001). Sole atony occurred less frequently than expected (8.2% in non-severe and 35.5% in severe PPH, p<.001). FXIII showed the largest decrease of coagulation factors by far, from no (–12%) to non-severe (–20%) and severe postpartum hemorrhage (–32%, p<.001). Duration of the second stage of labor was longer in women developing postpartum hemorrhage later on (71 vs. 46 minutes, p=.004), but was not different between women with or without assumed main reasons.

**Conclusion:** Uterine atony frequency is low in non-severe postpartum hemorrhage, but progresses from non-severe to severe postpartum hemorrhage. It can thus not be the frequent reason for postpartum hemorrhage it is assumed to be, as all postpartum hemorrhages start as non-severe. A prolonged second stage of labor together with an ongoing (likely self-reinforcing) consumptive coagulopathy helps to explain postpartum hemorrhage genesis. FXIII is a prepartum predictor of postpartum blood loss and shows the most pronounced peripartum coagulation factor loss in any setting. This might allow to identify new treatment pathways.

## INTRODUCTION

WHO defines PPH as a blood loss of ≥ 500 ml within 24 hours postpartum (> 1000 ml being considered major or severe). PPH still causes up to 1/3 of all maternal deaths, with 8-10 women dying from PPH every hour^1^. While being a major burden in low-income countries, increasing PPH-related complications are observed in industrialized countries as well. Scottish data suggest a rise in major obstetric hemorrhages within a decade^2^; and the incidence of severe PPH and maternal mortality has significantly risen in the US until 2014^3, 4^.

As a large proportion of women experiencing severe PPH have – so far – no or few identifiable prepartum risk factors^1^, it is important to better understand PPH pathophysiology. We identified prepartum FXIII activity^5^ (FXIII) and the prepartum platelet count^6^ (PLT) to be associated with postpartum blood loss, confirming other such data^7^. Several studies in the perioperative, postoperative and trauma setting have suggested that loss of coagulation factors is very prevalent and relevant for the respective clinical course, management and outcome^8, 9^. We thus postulated that coagulation factors might not only be relevant for the outcome, but also for the development of PPH.

While various risk factors for PPH have been described^10^, the current clinical doctrine suggests that essentially all PPH cases emerge from uterine atony (∼70% of cases), lacerations (∼20%) or placenta-related problems (such as retained placenta or placental tissue and placenta accreta spectrum, ∼10%)^11^. Thus, to better understand PPH development, our above findings (on FXIII and PLT) need to be put in perspective with these assumed main reasons (AMRs) for PPH. Coagulation factor changes over time from pre– to postpartum obviously describe the development of a PPH better than a single point measurement at the time of PPH detection. This follows the idea of detecting “the departure from the healthy state” rather than “detecting the onset of disease”, as outlined by Mitrea and coworkers^12^. We therefore evaluated the peripartum course of the newly identified prepartum markers of postpartum blood loss (FXIII and PLT), as well as the levels of fibrinogen and F II activity; PPH evolution (in it’s different severities) according to blood loss; and the presence or absence of AMRs for PPH. As clinical prepartum properties (e.g. BMI, duration of the second stage of labor) can also be potential departure points from a healthy state^13^, we investigated them in the context of postpartum blood loss as well.

## METHODS

### Enrollment and data collection

The PPH1300 study (a prospective, monocentric cohort study at the Department of Obstetrics, University Hospital Zurich (USZ), Switzerland) was approved by the local ethics review board (KEK-ZH 2015-0011) and registered with ClinicalTrials.gov (NCT02604602). Eligible women (≥18 years, gestational age ≥22 weeks, admission to the labour and delivery ward) were enrolled consecutively; women with known congenital coagulopathies were not enrolled. Women gave written informed consent. Clinical data were prospectively collected by dedicated research assistants, using IntelliSpace Perinatal (Philips, USA), Perinat (proprietary USZ database), and KISIM (cistec AG, Switzerland) software. Only women with vaginal deliveries were included in this study, as pathophysiology of PPH after caesarean section might be different from the one after vaginal delivery^14^.

### Blood collection, sample processing

Upon occurrence of regular contractions or rupture of membranes, the first blood draw was performed at admission for delivery. A second blood draw was performed 24 to 48 hours after delivery. In both instances, blood from an antecubital vein was collected into Vacutainer® tubes (6.0 mL, 9:1 diluted with 0.105 M buffered Na-citrate) and was processed without delay (centrifugation at 2800g and 17°C for 10 minutes; supernatant was carefully aliquoted, frozen and kept at –80°C) on a 24/7 basis. Care was taken to keep the samples frozen on dry ice during transportation to the study laboratory (Centre for Laboratory Medicine St. Gallen). Hemoglobin concentration and platelet counts were determined at the point of care.

### Measured blood loss (MBL)

Blood loss measurements were performed using a validated, quantitative measurement system graduated in 0.1 L steps, which shows excellent correlation with calculated blood loss in vaginal deliveries^15^.

### Assumed main reasons (AMRs) for PPH

Uterine atony was diagnosed by palpation of a flaccid and large uterus postpartum, despite the appropriate use of routine uterotonics.^16^ Lacerations were identified through careful inspection. The placenta was deemed retained if not expelled within 30 min postpartum with adequate traction on the umbilical cord. Retained placenta tissue was identified by inspection of the placenta and / or ultrasonography. No women with placenta accreta spectrum disorder were included in this study.

### Measurements of coagulation factors, platelet count and hemoglobin

Samples were collected at the beginning of the birth process; and 24-48 h after delivery; thus, all changes occurring during the WHO defined interval (up to 24 h post partum) were captured. Fibrinogen (FIB, Clauss method, Werfen), D-Dimer (immunological assay, Werfen) and FII activity (FII, clotting assay, Werfen) were determined on an ACLtop 750 (Werfen), FXIII activity (chromogenic assay, Siemens) on a XP (Siemens) analyzer. Hemoglobin concentrations (HGB) and PLT were analyzed on an ADVIA (Siemens) or a XN-20 (Sysmex). All assays were performed according to the recommendations of the manufacturers.

Women and caregivers were blinded to the results as samples were analyzed only after study recruitment was completed.

### Strata definition and statistical procedures used

Women were stratified according to the measured blood volume lost (MBL) (< 500 ml; ≥ 500 ml and < 1000 ml; ≥ 1000 ml: no, non-severe or severe PPH), the presence or absence of AMRs was observed. The ratio of post– to prepartum HGB was used as a marker of plausibility for measured peripartal blood loss, in combination likely to be the most adequate reflection of blood loss in PPH^17^.

### Differences of observed vs expected frequencies of AMRs were evaluated by a Chi-square test

Peripartum changes of HGB, PLT, FIB, FII and FXIII were described by the ratio of the post– to the prepartum results for each woman. Results were then compared according to stratification (groups with no, non-severe or severe PPH) as well as groups with presence or absence of AMRs. Results are presented as median with interquartile ranges [IQR]. Comparisons for statistically significant differences were done by Friedman, Mann-Whitney or Kruskal-Wallis testing, as appropriate.

The influence of the length of the second stage of labor (DUR), parity, age, gestation age and BMI on MBL was evaluated by continuous outcome logistic regression analysis.^18^ Influences of the pre– to postpartum changes in PLT, FIB, FII and FXIII on MBL were similarly evaluated. These parameters were also compared between women with and without AMRs.

Alpha levels of .05 or less were considered to identify statistical significance. Statistical analyses were independently performed with MedCalc® V23.0.5 and R (4.4.3, https://www.R-project.org/). All authors had access to the primary clinical trial data.

## RESULTS

Overall, 1500 women were recruited for the study. In 133 women, blood draws were not performed in time according to the study protocol; and tubes collected from 58 women were not adequately filled. Thus, data from 1309 women were evaluable. Of these, one woman (who did not develop PPH) was excluded after detection of a dilution error; thus 676 women who had vaginal deliveries represent the study population described here. They were further stratified according to the occurrence and degree of PPH; and whether or not AMRs for PPH were present (see study flow chart, figure 1). Age, BMI, gestation age, parity, duration of second stage of labor and prepartum HGB, FIB, FII and FXIII were not different between those with or without AMRs; only PLT showed a difference (189 G/l [165 – 219] vs. 205 G/l [173 – 245], p=0.006).

**Figure 1:**
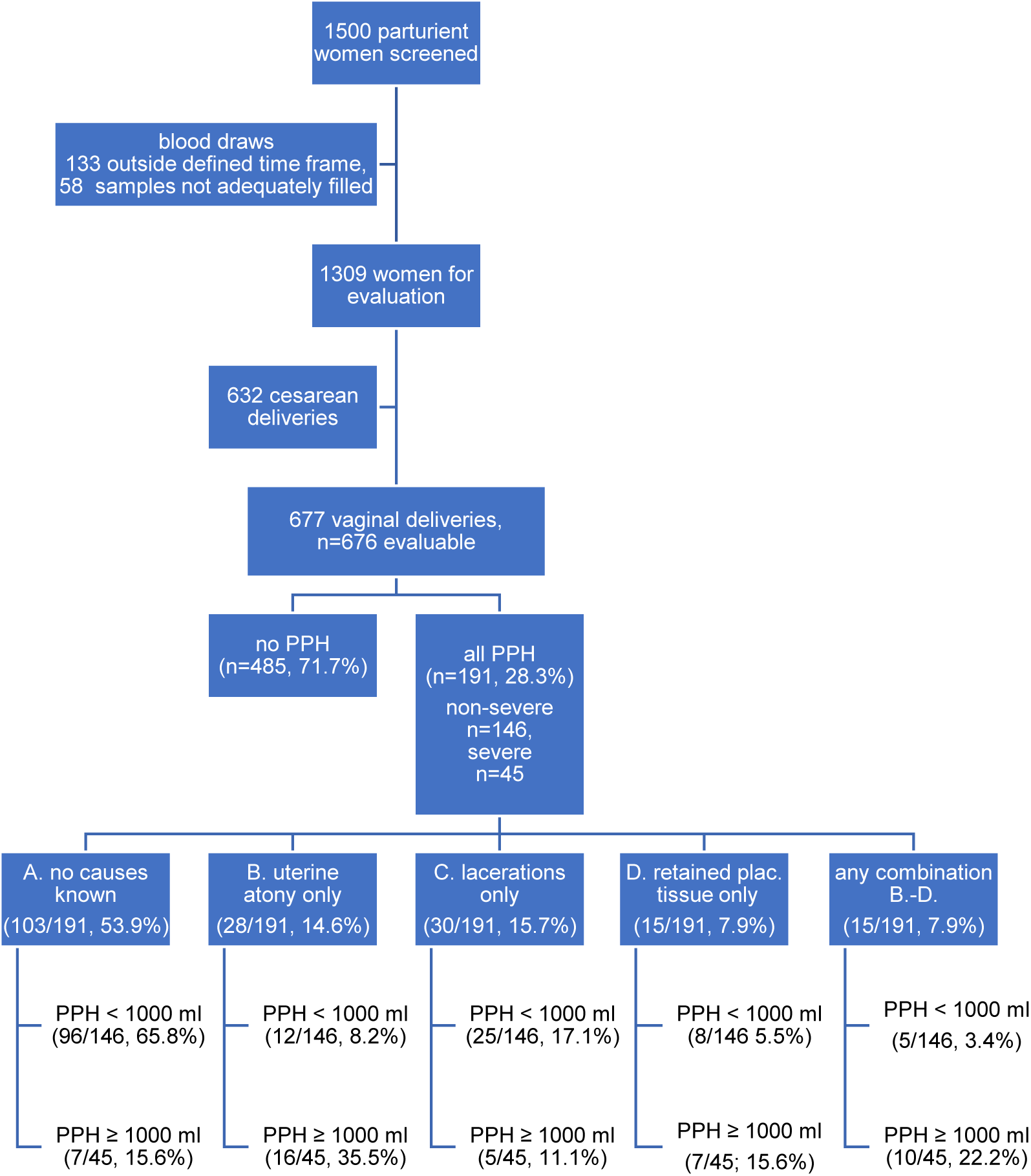
The numbers of patients excluded, included, evaluable and stratified patients and the numbers of patients with AMRs in the PPH stratum are displayed.

The results of HGB ratios (post– to prepartum levels) in the different PPH strata are displayed in Table 1. The significant differences between the different strata (p<0.001) are well compatible with the observed MBL, speaking against misclassification and validating the ratio between post– and prepartum result as an adequate description of the peripartum course.

**Table 1:** Course of hemoglobin concentration from pre– to postpartum (as median ratio: post-/prepartum result) in the different PPH strata (no PPP (MBL < 500 ml), non-severe PPH (≥ 500 ml, < 1000 ml), severe PPH (≥1000 ml)): the observations made by MBL to categorize PPH are plausible.

|  | **no PPH** | **Non-severe PPH** | **Severe PPH** |
| --- | --- | --- | --- |
| HGB, peripartum course, median [IQR] | 0·91 [0·87 – 0·97] | 0·84 [0·78 – 0·89] | 0·75 [0·66 – 0·79] |
| Loss of pre- to postpartum HGB | 9% | 16% | 25% |
| MBL, median [IQR], ml | 300 [250–400] | 600 [500–738] | 1200 [1150–1500] |

Of the 676 women, 191 developed PPH. This was non-severe in 146 women, of which 96 were without any of the traditionally known AMRs for PPH (without AMRs observed 65.8% vs. expected <1%, p<.001); 12 women showed uterine atony (8.2% vs. expected 70%, p<.001), 25 women had lacerations (17.1% vs. 20%, p=.63), 8 women retained placental tissue (5.5% vs. 10%, p=.31), and 5 women (3.4%) showed any combination of AMRs.

Of 45 women with severe PPH, seven were without any AMRs (observed 15.6% vs. expected <1%, p<.001), 16 women had uterine atony (35.6% vs. 70%, p<.001), 5 women had lacerations (11.1% vs. 20%, p=.14), 7 women retained placental tissue (15.6% vs. 10%, p=.21), and 10 (22.2%) any combination of AMRs.

Even if all PPH cases (non-severe and severe) with “uterine atony” and “any combination of AMRs” were summoned as “uterine atony”, the observed frequency remained far below the traditionally expected frequency for uterine atony in PPH (22.5% observed vs. 70% expected, p<.001). For an overview, see Figure 1.

The peripartum changes of PLT, FIB, FII and FXIII within the different degrees of PPH and the comparison for statistically significant differences between these groups are presented in Table 2.

**Table 2:**
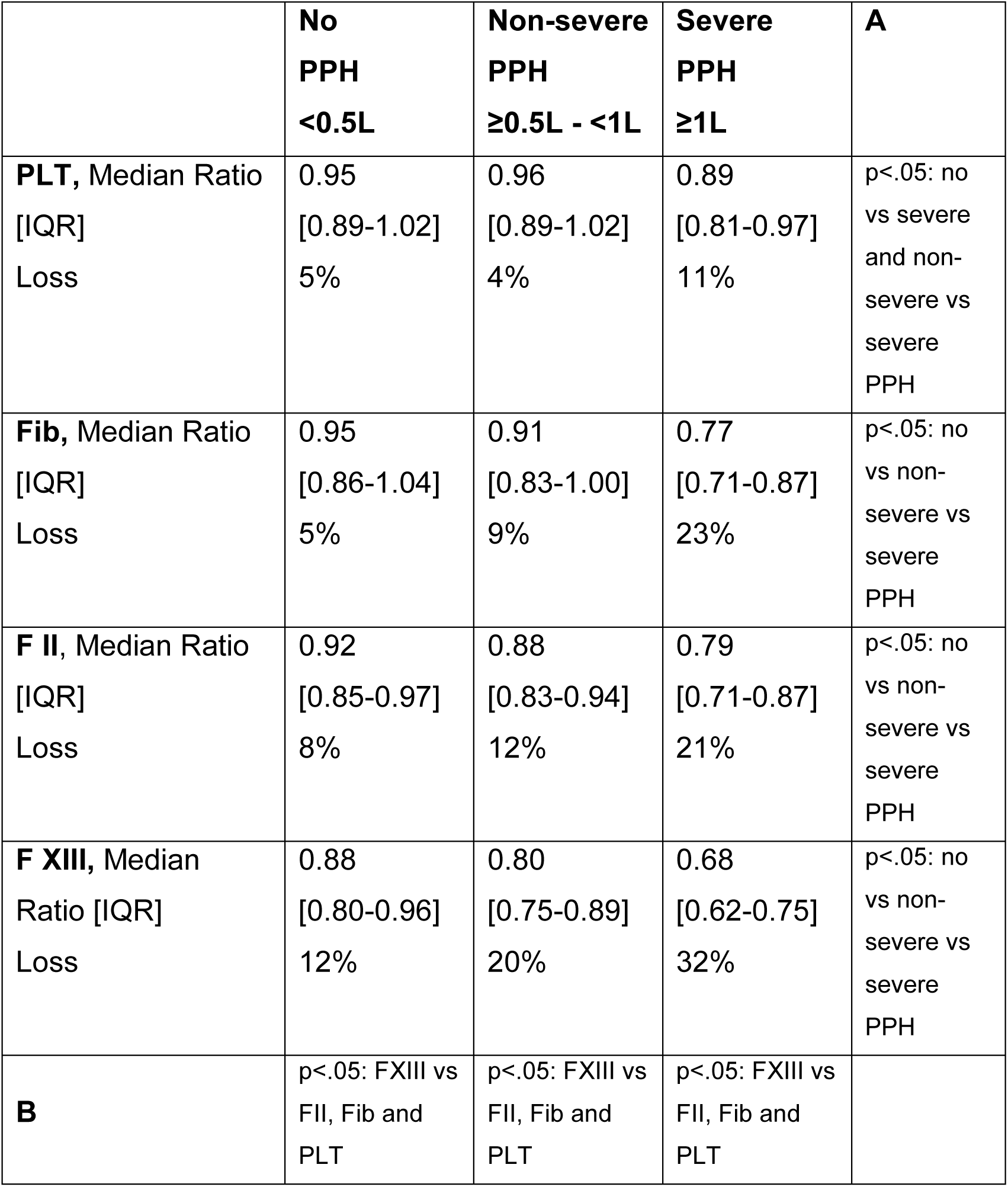
Comparisons of loss of coagulation factors (from pre– to postpartum, median ratio [interquartile ranges]) in 676 women with vaginal deliveries: a) for the respective coagulation factor between the different PPH strata (no, non-severe or severe PPH), statistical results are given in colum “A”. b) between the different coagulation factors (PLT, Fib, FII, FXIII) within the same PPH stratum; statistical results are given in row “B”.

Results of peripartum changes for PLT, FIB, FII and FXIII within the different degrees of PPH and the comparison for statistically significant differences between the respective groups of women with or without AMRs are presented in table 3. No relevant differences were observed between women with and without AMRs, a graphical display for FXIII is presented in Figure 2.

**Figure 2:**
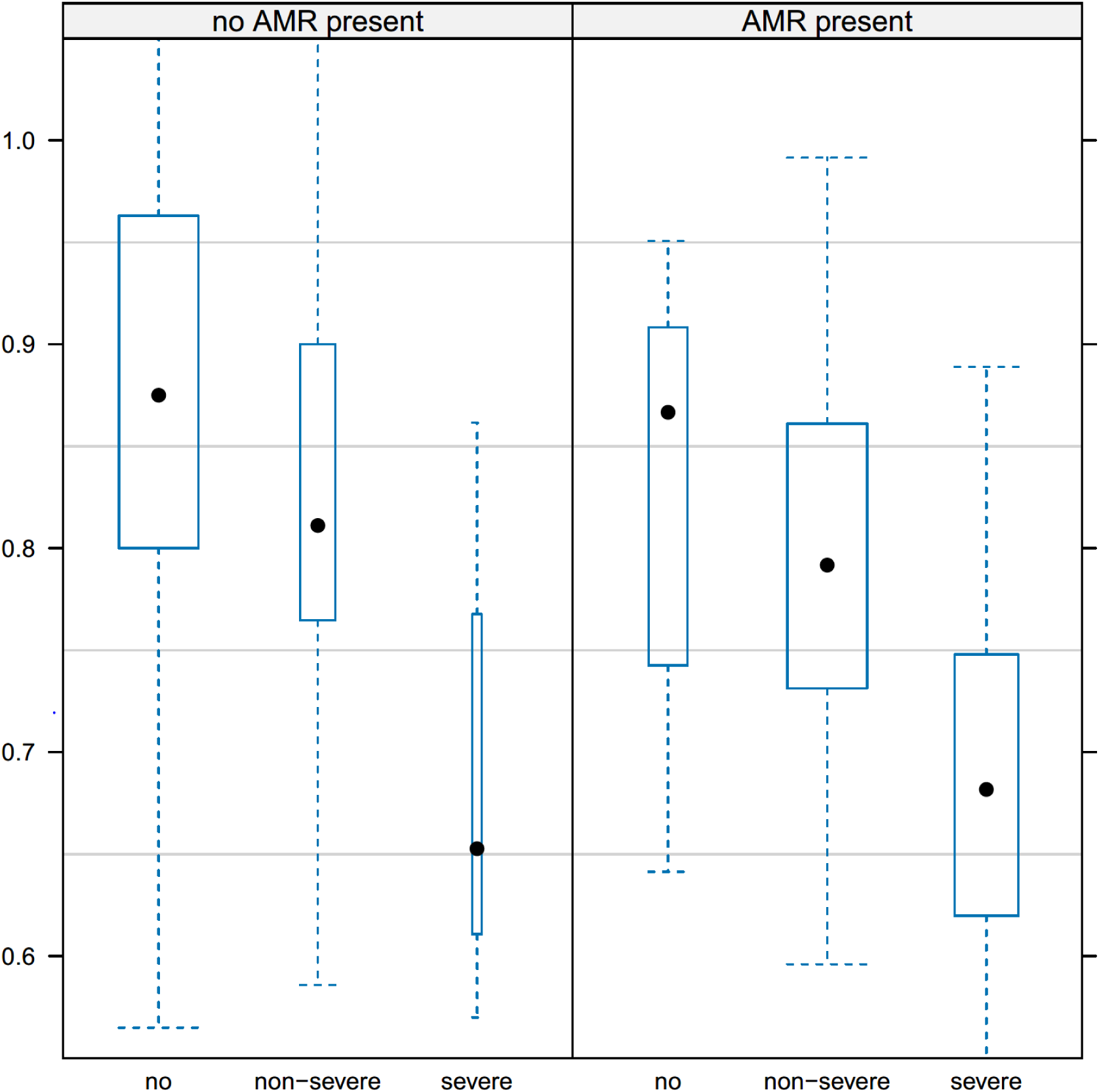
Comparison of the peripartum course of FXIII (post-/prepartum ratio, median, IQR) in women without and with assumed main reasons (AMR) for PPH. The similar progressive decrease from no to non-severe and from non-severe to severe PPH, independent of the presence or absence of AMRs for PPH, can be seen.

**Table 3:** The loss of coagulation factors from pre– to postpartum (median ratio [interquartile range]) in 676 women with vaginal deliveries with “no PPH” (MBL < 500 ml), “non-severe PPH” (≥ 500 ml to < 1000 ml) and “severe PPH” (≥ 1000 ml) in the strata “assumed main reasons” and “no assumed main reasons” are shown; as well as the level of statistical significance for the differences between the strata.

|  | **NO “assumed main reasons” for PPH present** | **“Assumed main reasons” for PPH present** | **p =** |
| --- | --- | --- | --- |
| **Platelets** |  |  |  |
| No PPH Loss | 0.95 [0.89-1.02]<br>5% | 0.95 [0.89-0.97]<br>5% | .45 |
| Non-severe PPH Loss | 0.97 [0.90-1.02]<br>3% | 0.95 [0.88-1.02]<br>5% | .26 |
| Severe PPH Loss | 0.84 [0.82-0.88]<br>16% | 0.90 [0.80-0.98]<br>10% | .20 |
| **Fibrinogen** |  |  |  |
| No PPH Loss | 0.95 [0.86-1.04]<br>5% | 0.89 [0.85-1.02]<br>11% | .51 |
| Non-severe PPH Loss | 0.92 [0.84-1.00]<br>8% | 0.86 [0.82-0.99]<br>14% | .04 |
| Severe PPH Loss | 0.76 [0.69-0.81]<br>24% | 0.80 [0.71-0.87]<br>20% | .42 |
| **F II** |  |  |  |
| No PPH Loss | 0.92 [0.85-0.97]<br>8% | 0.88 [0.79-0.91]<br>12% | .09 |
| Non-severe PPH Loss | 0.89 [0.83-0.94]<br>11% | 0.87 [0.82-0.93]<br>13% | .26 |
| Severe PPH Loss | 0.74 [0.64-0.82]<br>26% | 0.81 [0.72-0.87]<br>19% | .23 |
| **F XIII** |  |  |  |
| No PPH Loss | 0.88 [0.80-0.96]<br>12% | 0.87 [0.76-0.91]<br>13% | .24 |
| Non-severe PPH Loss | 0.81 [0.76-0.90]<br>19% | 0.79 [0.73-0.86]<br>21% | .11 |
| Severe PPH Loss | 0.65 [0.61-0.77]<br>35% | 0.68 [0.62-0.75]<br>32% | .75 |

Continuous outcome logistic regression analysis revealed that DUR was a relevant explanatory variable associated with the extent of MBL (odds ratio 0.80, 95% confidence interval 0.70 – 0.92, for every 60 minutes not spent in labor, p=.001) even after correction for other clinical variables such as age, BMI, gestation age and parity. Gestation age (p=.07) and parity (p=.09) showed strong trends to influence MBL.

DUR was significantly longer in women who later developed PPH (median 46 [17-114] without vs. 71 [22-140] minutes with PPH, p=.004); no difference in DUR was seen in women with or without AMRs (median 57 vs. 51 minutes, p=.32). Prepartum D-Dimer concentrations were elevated in women who later developed PPH (1.54 mg/l [1.24 – 2.01] without vs 1.66 mg/l [1.34 – 2.32] with PPH, p=.008), particularly those who developed severe PPH (2.07 mg/l [1.35 – 4.02]).

Continuous outcome logistic regression analysis also revealed that peripartum change of FXIII change was the only one significantly associated with the extent of MBL (OR 1.56, 95% confidence interval 1.35 – 1.80 for every 10% decrease, p<.001, after correction for changes in PLT, FIB and FII), and with FII showing a strong trend (p=.06).

### Comment

This study in 676 evaluable women with vaginal deliveries is, to our knowledge, the largest study so far to prospectively evaluate changes in various coagulation factors during delivery, the presence or absence of AMRs for PPH and various clinical factors.

Having identified the influence of prepartum FXIII^5^ and PLT^6^ on postpartum blood loss, we were eager to understand how these observations can be brought into congruence with the traditional concept of AMRs for PPH, i.e. uterine atony (believed to explain ∼70% of all cases), lacerations (∼20%) and placenta related problems (∼10%), which all occur intra– or postpartum.

### Principal Findings

While we observed lacerations and placenta related problems to occur with the expected frequencies, we were surprised to find that this was by far not the case for uterine atony, both in patients with non-severe and severe PPH. More than half of the PPH cases could not be explained by the “traditional” AMRs, in particular by uterine atony (see Figure 1). At the same time, we documented that patients with PPH show a similar consumption process of the coagulation system, be it with or without the presence of AMRs (see table 3); and that F XIII is the coagulation factor most sensitive to changes occurring during delivery (see table 2).

### Results in the Context of What is Known

A lower than supposed frequency of uterine atony in PPH has been described before by other research groups (see table 4). However, these results seem not to have triggered a search for other explanations of PPH genesis so far, possibly due to the fact that in unclear cases, “The majority of cases are traditionally attributed to atony.”^19^ The low proportion of women with sole uterine atony in non-severe (8.2%) and severe PPH (35.5%), casts doubt on the idea that uterine atony is the most prevalent cause for PPH. This observation is contrary to current doctrine and challenges the assumed causality of uterine atony for any PPH – as every severe PPH obviously transits a phase of non-severe PPH to begin with. Thus, including observations from other research groups, these findings seem to suggest that uterine atony cannot be the frequent cause for PPH it is traditionally believed to be.

**Table 4:**
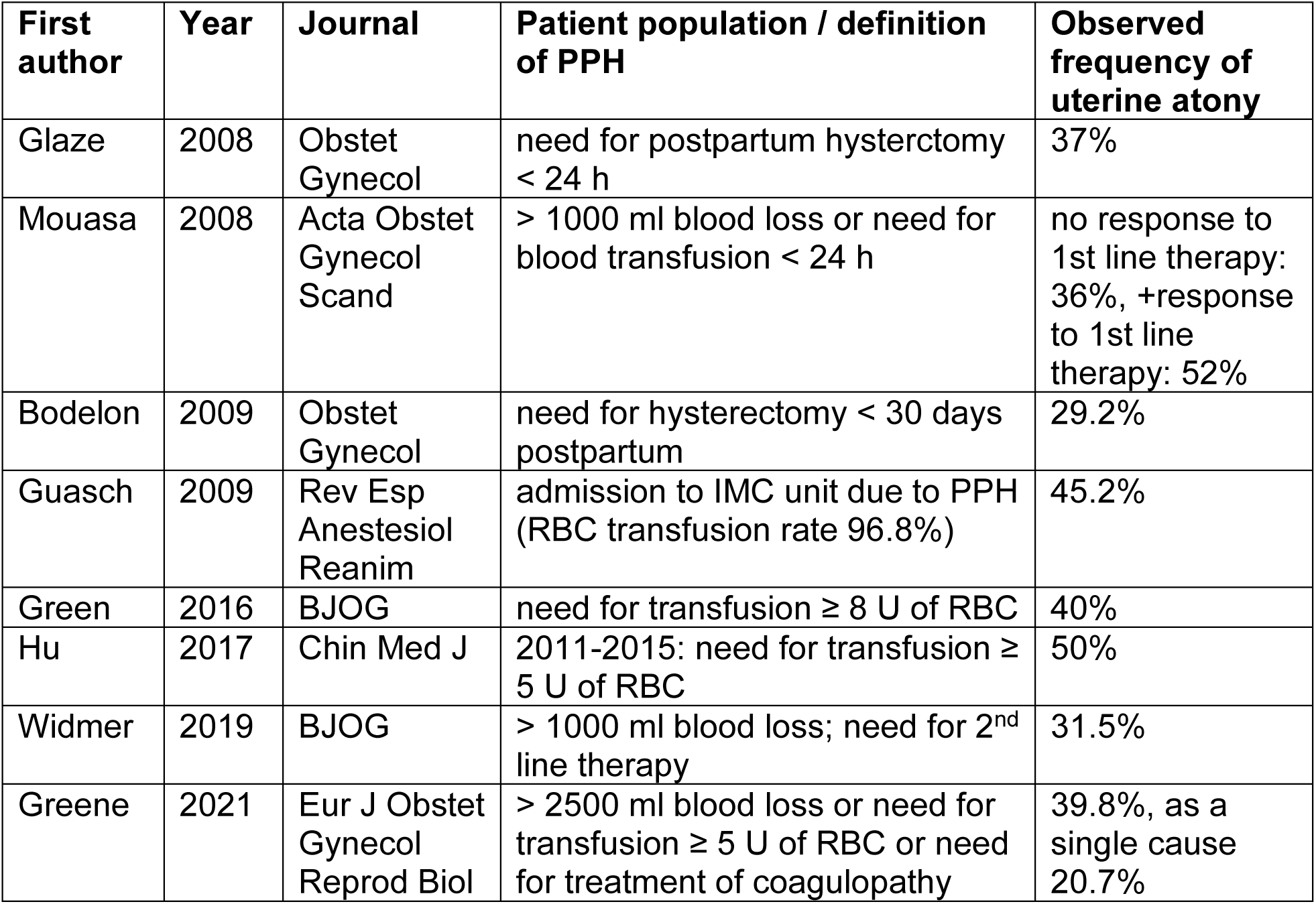
Results of other published results with regard to the proportion of uterine atony observed in women with PPH are displayed.

We therefore evaluated the influence of the presence or absence of AMRs and PPH on the peripartum courses of PLT, FIB, FII and FXIII: results were similar in patients with or without AMRs, both in non-severe and severe PPH (table 3, exemplary figure 2 for FXIII). This is further support to the idea that “traditional” AMRs cannot explain a very large proportion of PPH occurrences.

The extent of changes in coagulation factors in women without PPH, non-severe or severe PPH was different between PLT, FIB, FII and FXIII. PLT and FIB seem the least sensitive components to loss (see tables 2, 3). This might be explained by the respective prepartum course: pregnancy at term comes with increased megakaryocytic stimulation through TPO^20^, which increases platelet life span and platelet mass^21^. And FIB steadily increases during gravidity, leading to increased levels at term^22^, while FII is essentially unchanged. On the other hand, FXIII steadily decreases towards delivery^23, 24^, indicating that synthesis does not compensate for the pregnancy associated pre– (and peri-) partum decrease. Observations from others and ourselves^25–27^ let us assume that, compared to the other factors evaluated (and in line with our earlier results^5^), the most pronounced susceptibility of FXIII to peripartum loss and/or consumption observed here likely renders the final pathway vulnerable to increasing clot instability^28^.

The question then remains what might stimulate the development of PPH (and specifically, in those women who do not have AMRs). The duration of the second stage of labor has been associated with the risk of developing PPH^29^; and preexisting or worsening coagulopathies seemingly support PPH^30^. In the analyses described above, we found that DUR, after correction for age, parity, gestation age and BMI, is significantly influencing the amount of blood lost during delivery. While DUR was shown to be not different between patients that did or did not show AMRs later on, it was significantly prolonged in patients that were going to develop PPH (vs no PPH) later on. The coagulation system in patients who later develop PPH is sufficiently activated to support consumption as evidenced by the moderately, but significantly increased D-dimer concentration at admission for delivery. These observations allow us to suggest that a sufficiently activated coagulation system during a prolonged second stage of labor may induce or support the development of a PPH, associated with a continued consumption of coagulation factors (especially F XIII), even if AMRs are not present. The presence of a potentially ongoing consumptive process observed later on does not preclude that in addition, the coagulopathy of PPH might also harbor specific other changes^31^, with or without the presence of AMRs. Significant differences observed in the peripartum course of PLT, FIB, FII and FXIII (table 2) suggest that the (physiological) compensatory need is different between the various components, especially when it comes to FXIII.

It seems not too surprising that FIB proved to be a very robust coagulation factor with regard to peripartum changes, as it is not a marker of postpartum blood loss^22, 32^; this might help to explain why its use did not improve or prevent PPH in various randomized controlled trials^33–36^.

### Clinical and Research Implications

Our data imply that the evolvement of a PPH is more frequently (than assumed until now) associated with a developing, consumptive – and in the end coagulopathic – process, stimulated by a prolonged duration of the second stage of labor. This suggests that early replacement of most critical coagulation components (according to our data, this is F XIII) might attenuate or even preclude the development of a (severe) PPH. A publicly funded, prospective, controlled, randomized trial to elucidate this question is currently underway^37^.

### Strengths and limitations

Strengths of our study are the prospective approach in a “real world” setting, the large database, the homogeneous study performance, the measurement of postpartum blood loss (rather than estimating it) and the dynamic (pre/post) observation of the coagulation system in all participants (no, non-severe and severe PPH cases). The main limitation is that the verification of our postulate requires a prospective randomized clinical trial.

### Conclusion

We suggest that uterine atony, solely or in combination with other AMRs, occurs significantly less frequently than traditionally assumed, both in non-severe and severe PPH. Uterine atony is, therefore, unlikely to be the frequent cause for PPH it is usually believed to be. This applies particularly to patients with non-severe PPH, where atony seems to be frequently diagnosed due to the absence of other, more convincing causes; and in order to remain within the current medical doctrine.

Observations on the overall peripartum course of coagulation components show that a prolonged second stage of labor and acquired coagulation changes, particularly of FXIII, are stringently associated with the genesis of PPH. This helps to explain the development of PPH, independently of the presence or absence of AMRs, particularly uterine atony. However, this does not negate the possibility that PPH can develop due to AMRs if they are present.

In the light of our earlier results on FXIII as a prepartum predictor of postpartum blood loss, more research is needed to further elucidate the mechanisms involved. This approach might harbor the potential to identify coagulation components that can be used to treat – and hopefully one day prevent – a significant number of (severe) PPH cases.

## Data Availability

For original data, please contact. Deidentified individual participant data that underlie the reported results will be made available upon justified request.

## ACKNOWLEDGEMENTS

We would like to thank all participants for their support and effort. We thank all midwives, nurses and doctors involved; and Rita Bischof and Isabelle Kamm for their expert assistance in performing the laboratory analyses.

## AUTHORS CONTRIBUTIONS

W. Korte and C. Haslinger planned the study, data evaluation and drafted the manuscript. J. Bürgi and W. Korte oversaw the laboratory work. C. Haslinger and N. Ochsenbein oversaw the clinical data collection. W. Korte and M. Rösslein did statistical work using MedCalc, T. Hothorn independently did statistical work using “R”. W. Korte, C. Haslinger and T. Hothorn revised the manuscript. All authors approved the manuscript after final revision.

